# DNR orders in SARS-CoV-2 patients: a retrospective validation study in a Swiss COVID-19 Center

**DOI:** 10.1101/2021.07.12.21260359

**Authors:** Giorgia Lo Presti, Maira Biggiogero, Andrea Glotta, Carola Biondi, Zsofia Horvath, Rosambra Leo, Giovanni Bona, Alessandra Franzetti-Pellanda, Andrea Saporito, Samuele Ceruti, Xavier Capdevila

**Affiliations:** Clinical Research Unit, Clinica Luganese Moncucco, Lugano, Switzerland; Department of Critical Care, Clinica Luganese Moncucco, Lugano, Switzerland; Service of Internal Medicine, Clinica Luganese Moncucco, Lugano, Switzerland; Service of Radiotherapy, Clinica Luganese Moncucco, Lugano, Switzerland; Department of Anaesthesiology, Bellinzona Regional Hospital, Ente Ospedaliero Cantonale, Bellinzona, Switzerland; Department of Anaesthesia and Intensive Care, Montpellier University Hospital, Montpellier, France

**Keywords:** Palliative Care, COVID-19, SARS-COV-2, dyspnea, Do-Not-Resuscitate order

## Abstract

**Background:** The worldwide pandemic situation forced many hospitals to adapt COVID-19 management strategies. In this scenario, the *Swiss Academy of Medical Sciences* (SAMW/ASSM) organized national guidelines based on expert opinion to identify Do-Not-Resuscitate (DNR) patients, to reduce futile ICU admission and resource misuse. However, the practical impact of this standardized national protocol has not been yet evaluated. In our specialized COVID-19 Center, we investigated characteristics and mortality of DNR patients identified according to national standardized protocol, comparing them to non-DNR patients.

**Methods:** This was a pilot retrospective validation study, evaluating consecutive hospital admitted COVID-19 patients. Primary outcome was 30-days survival of *DNR patients* in comparison to the *control group*. Secondary outcomes reported quality of treatment of deceased patients, especially of agitation/sedation and dyspnea, using respectively the Richmond Agitation-Sedation Scale – Palliative care (RASS-PAL) for sedation and agitation (+4/-5) and the modified Borg Scale for dyspnea (1-10).

**Results:** From March 16 to April 1, 2020, 213 consecutive patients were triaged; at 30-days follow-up, 9 patients (22.5%) died in the *DNR group*, 4 (2.2%) in the *control group*. The higher mortality rate in the *DNR group* was further confirmed by Log-Rank Mantel-Cox (23.104, p < 0.0001). In the *DNR-group* deceased patients, end-of-life support was performed with oxygen (100%), opioids (100%) and sedatives (89%); the mean RASS-PAL improved from 2.2 to -1.8 (p < 0.0001) and the Borg scale improved from 5.7 to 4.7 (p = 0.581).

**Conclusion:** A national standardized protocol identified patients at higher risk of short-term death. Although the legal status of DNRs varies from country to country, the implementation of national standardized protocol could be the way to guarantee a better treatment of COVID-19 patients in a pandemic situation with limited resources.

## INTRODUCTION

During the beginning of 2020, the world witnessed the first COVID-19 pandemic^1^. This dramatic situation forced many hospitals to improve their competence and in some countries such as in Switzerland some hospitals became dedicated centers to provide COVID-19 patients’ specific management, following governmental directions ^2^.

In this pandemic scenario, the management of COVID-19 patients at high risk of morbidity and mortality raises new aspects of concerns, due to the risk of futilely investing precious resources such as CPR or ICU-level care^3^. As a consequence, Health-Care Professionals (HCP) had to find ways to identify those patients to whom the application of resources related to ICU admission resulted useless, especially when faced with limited hospitalization capacity and with the potential risk of shifting resources and time away from patients who could benefit from them. The Do-Not-Resuscitate (DNR) order is applied to those patients for whom further medical treatment is considered to be futile, with a very high probability of death or ventilator dependency despite the use of additional therapies^4,5^. A careful management of resources was also applied during the H1N1 influenza pandemic, in 2009, when the UK Resuscitation Council suggested an early identification of DNR patients, in order to avoid futile resource wasting^6^ and thus allowing a reduction in ICU hospitalizations^7,8^.

The legal status of DNRs varies from country to country^5^, the Switzerland experience is connected to the role of the Swiss Academy of Medical Sciences (SAMW/ASSM), a bridge builder between science and society, which implemented and applied a national standardized protocol of care based on expert opinion, with the aim to guarantee a better treatment of COVID-19 patients in an emergency situation with limited resources ^9,10^. The protocol determined the indications for a correct evaluation and identification of patients suitable for a DNR order. This protocol supported HCP decision at triage and at hospital admission, with the aim to identify patients at high risk of death despite a potential ICU support, thus avoiding futile treatments, ICU overload and additional health costs ^3,9^. Nevertheless, until today, the practical consequences of the SAMW/ASSM guidelines are still unknown.

According to other authors, DNR order resulted as an independent risk factor for death in the post-operative period, with DNR patients presenting higher fatality rate^11^, a more rapid disease progression and a more frequent need of an early Palliative Care (PC) ^12,13^, especially to improve the treatment of dyspnea, restlessness, anxiety and delirium ^14^. The early identification of DNR patients requiring PC treatment in the short-term during hospital stay is therefore mandatory for a better patients’ management and also from an ethical point of view, this last point acquiring even more importance in the current pandemic scenario.

Aim of this study was to report and analyze the survival rate of DNR COVID-19 patients hospitalized in a single Swiss COVID-19 center, identified and managed as DNR according to the SAMW/ASSM guidelines, to retrospectively validated the effectiveness of the Swiss national standard protocol in an emergency pandemics situation characterized by scarce availability of resources.

## METHODS

This was a pilot retrospective validation study, evaluating consecutive DNR patients hospitalized with a diagnosis of COVID-19 during the first two weeks of the first pandemic wave. DNR order was defined according to the first version of SAMW/ASSM national guidelines (Table 1) ^9^, defining patients ineligible for ICU admission when they presented, between others, severe cerebral deficits after a stroke, NYHA class III or IV heart failure, COPD GOLD IV, liver cirrhosis with refractory ascites or encephalopathy at stage > I, stage V chronic kidney disease, a Frailty index higher than 4, age greater than 80 years and/or an estimated survival of less than 24 months ^9^.

**TABLE 1.** Criteria for ICU admission, according to SAMW/ASSM guidelines.

|  |
| --- |
| **Step 1: Does the patient have any of the following inclusion criteria?**<br>• Requirement for invasive ventilatory support?<br>• Requirement for hemodynamic support with vasoactive agents (noradrenaline-equivalent dose >0.1 µg/kg/min)?<br>If one of these inclusion criteria is fulfilled → Step 2 |
| **Step 2: Does the patient have any of the following exclusion criteria?**<br>• Patient's wishes (advance directive, etc.)<br>• Unwitnessed cardiac arrest, recurrent cardiac arrest, cardiac arrest with no return of spontaneous circulation<br>• Malignant disease with a life expectancy of less than 12 months<br>• End-stage neurodegenerative disease<br>• Severe and irreversible neurological event or condition<br>• Chronic conditions:<br>○ NYHA class IV heart failure<br>○ COPD GOLD 4 (D)<br>○ Liver cirrhosis, Child-Pugh score >8<br>○ Severe dementia<br>○ Severe circulatory failure, treatment-resistant despite increased vasoactive dose (hypotension and/or persistent inadequate organ perfusion)<br>• Estimated survival <12 months |

*Triage ICU admission criteria according to SAMW/ASSM guidelines, to maximize personal benefit with public health community resources.*

### Outcomes

Primary outcome was the 30-day overall survival in patients defined as DNR according to the SAMW/ASSM guidelines (*DNR group*), analyzing with validation tests whether this group of patients was at greater risk of death than patients defined as not-DNR (*control group)*. Secondary outcomes were to report the quality of symptoms management according to patients’ clinical status, reporting their clinical characteristics, with a special focus on dyspnea and on the degree of agitation/sedation.

### Information dataset

Through the health medical records, a dataset containing biological, clinical and medical information of DNR patients hospitalized during the study period was created. Biological data concerned age, sex, frailty status ^15^, comorbidities (oncological disease, heart failure, COPD, cirrhosis and neurodegenerative disease) and all characteristics regarding DNR application were collected. Finally, the 30-day mortality rate was also reported.

Clinical symptoms were evaluated using standard score systems; the most frequent symptoms encountered in COVID-19 patients were breathlessness and agitation ^16^. Dyspnea was evaluated according to the modified Borg scale ^17^, a semiquantitative numerical scale frequently used as measure of perceived exertion during physical activity. Sedation and agitation were evaluated according to the Richmond Agitation Sedation Scale modified for palliative care patients (RASS-PAL) ^18^. The modified Borg and the RASS-PAL scales were regularly evaluated twice a day by nursing staff; moreover, patients were further reassessed and revaluated with the above-mentioned scales whenever a variation in dyspnea or agitation level was registered. All medical relevant information concerning the most important pharmacological therapies (use of opioids, sedatives and neuroleptics), fluid therapy, oxygen delivery were also reported. Considering that a pandemic is a social health problem, family visit restriction was implemented for all patients due to Public Law 19, the only exception being a single 15-minute visit, performed by only one family member, to patients who entered end-of-life care approach.

### Statistical analysis

Descriptive statistics of frequency was reported. Categorical variables were expressed as number (percentage); continuous variables were expressed as average (standard deviation, SD). Data distribution was verified with Kolmogorov-Smirnov test and with Shapiro-Wilk test. To identify significant difference between continuous variables, t-tests were performed. A Chi-Square statistic with 1 degree of freedom was carried out to study differences between categorical variables. Kaplan-Meier analysis was used to study patients’ survival with the Cox-Mantel log-rank test to ascertain differences between the groups, analyzing all event by time to hospital admission. All confidence intervals (CI) were established at 99%; p-value significance level was established to be < 0.01. Statistical data analysis was performed using the SPSS.26 package (SPSS Inc., Armonk, NY; USA).

### Ethics considerations

The project has been approved by the local Ethics Committees (CE TI 3682) according to the local Federal rules. All the participants provided a written informed consent form.

## RESULTS

During the first two weeks of pandemics, from March 16 to April 1 2020, 242 consecutive patients were admitted to the Hospital; 29 patients were excluded, as they had already been directly admitted as critically ill patients to the ICU. Of 213 patients not-critically ill at hospital admission, 40 (18.8%) were identified by HCP as DNR patients according to SAMW/ASSM national guidelines, thus resulting ineligible for ICU admission (*DNR group*); 173 (81.2%) patients were instead admitted to the internal medicine Department without a DNR order (*control group*, Figure 1). Among the *DNR group*, 27 (67.5%) patients presented a Frailty Index greater than 4, 21 (52.5%) had at least one chronic disease, 7 (17.5%) had a diagnosis of severe circulatory failure and 6 (15%) were affected by oncological active or metastatic disease. Among the *DNR group*, mean age was 79.9 years (62 – 97, SD 7.9) while among the *control group* it was 63.2 years (24 – 86, SD 12.1, p = 0.011); male patients were 25 (64%) in the *DNR group*, and 103 (59.5%) in the *control group* (Table 2).

**Figure 1.**
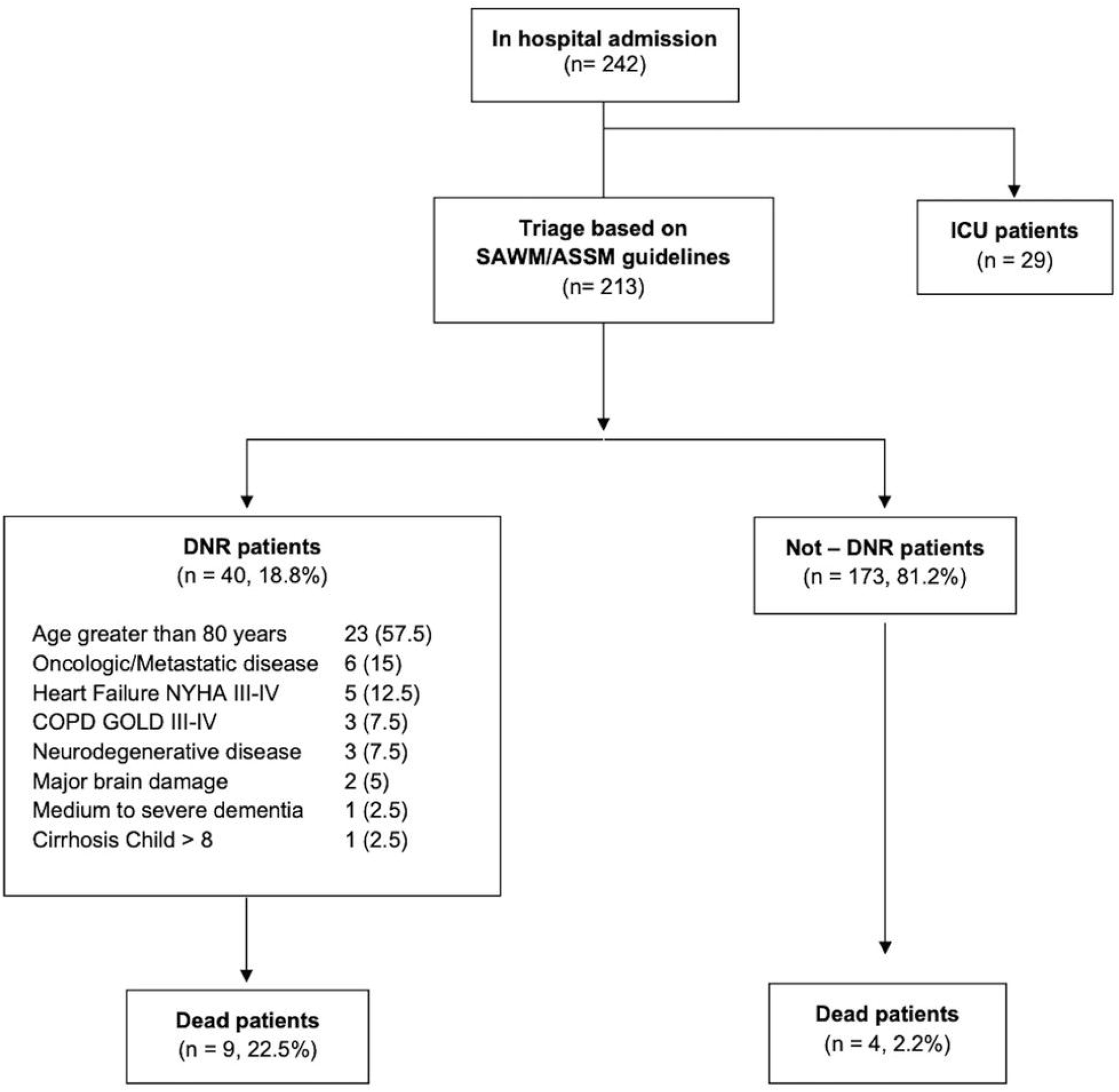
CLM COVID-19 patients’ stratification during first two weeks of pandemics, according to DNR order identified by SAMW/ASSM guidelines.

**TABLE 2.** Demographics of patients stratified according to SAMW/ASSM guidelines.

|  | **DNR group** | **Control group** | **p value** |
| --- | --- | --- | --- |
| **Number** | 40 (18.8) | 173 (81.2) |  |
| **Age** | 79.9 (62 – 97, SD 7.9) | 63.2 (24 – 86, SD 12.1) | 0.011 |
| **Sex male** | 25 (64) | 103 (59.5) | 0.257 |
| **Comorbidities** |  |  |  |
| **Frailty Index > 4** | 27 (67.5) | -- |  |
| **Age greater than 80 years** | 23 (57.5) | -- |  |
| **Oncologic/Metastatic disease** | 6 (15) | -- |  |
| **Heart Failure NYHA III-IV** | 5 (12.5) | -- |  |
| **COPD GOLD III-IV** | 3 (7.5) | -- |  |
| **Neurodegenerative disease** | 3 (7.5) | -- |  |
| **Major brain damage** | 2 (5) | -- |  |
| **Medium to severe dementia** | 1 (2.5) | -- |  |
| **Cirrhosis Child > 8** | 1 (2.5) | -- |  |
| **Outcome** |  |  |  |
| **Dead rate at 30 days** | 9 (22.5) | 4 (2.2) | < 0.001 |

*Demographics and medical information of DNR patients. Data are represented as means (min-max, SD) or medians (IQR) for non-normally distributed variables.*

### Primary Outcome

At 30-days follow-up, 9 (22.5%) patients in the *DNR group* had died due to cardiovascular arrest (Table 3); mortality rate in the *control group* was 2.2% with 4 patients deceased (Figure 1 and 2). Concerning the deceased patients in the *DNR group*, mean age was 82.4 years (72 – 93, SD 6.9), all of them lived at home before the hospitalization, 7 (78%) presented a Frailty index higher than 4, 3 (33%) a pre-existing severe heart disease, 2 (22%) a metastatic oncological disease and 1 (11%) a chronic brain damage (Figure 1). In order to perform a preliminary guidelines’ validation ^9^, a contingency table was structured according to the outcome stratification and *DNR group* allocation. A Chi-square test comparing mortality in the two groups, revealed a higher mortality rate in the *DNR group*, p < 0.0001 (Table 4SM); similarly, the Kaplan-Meier distribution reported a significant difference over time (Log-Rank Mantel-Cox 23.104, p < 0.0001, Figure 2).

**TABLE 3.** Demographic of deceased patients.

| **Deceased patients (n = 9)** |  |
| --- | --- |
| **Age** | 82.4 (72 – 93, SD 6.9) |
| **Male** | 7 (78) |
| **Comorbidities** |  |
| **Frailty Index >4** | 7 (78) |
| **Chronic disease $\geq 1$** | 6 (67) |
| **Severe heart failure** | 3 (33) |
| **Oncological active / metastatic disease** | 2 (22) |
| **Chronic brain damage** | 1 (11) |
| **Clinical information** |  |
| **Hospital LOS** | 6.2 (3 – 11, SD 2.4) |
| **RASS-PAL Before PC** | 2.2 (0 – 4, SD 1.2) |
| **RASS-PAL After PC** | -2 (-3.5 – -0.5) |
| **Borg Scale Before PC** | 6 (3.2 – 8) |
| **Borg Scale After PC** | 4.7 (0 – 10, SD 3.7) |
| **Medical therapies** |  |
| **Opioids** | 9 (100) |
| **Oxygen delivery** | 9 (100) |
| **Sedatives** | 8 (89) |
| **Antimicrobial/anti-retroviral therapy** | 8 (89) |
| **Fluid therapy** | 4 (44) |
| **Neuroleptics** | 0 |
| **Other** |  |
| **Family members visit before death** | 8 (89) |

*Demographics and medical information in deceased patients' subgroup of DNR patients. Data are presented as means (min-max, SD) or medians (IQR) for non-normally distributed variables.*

**Figure 2.**
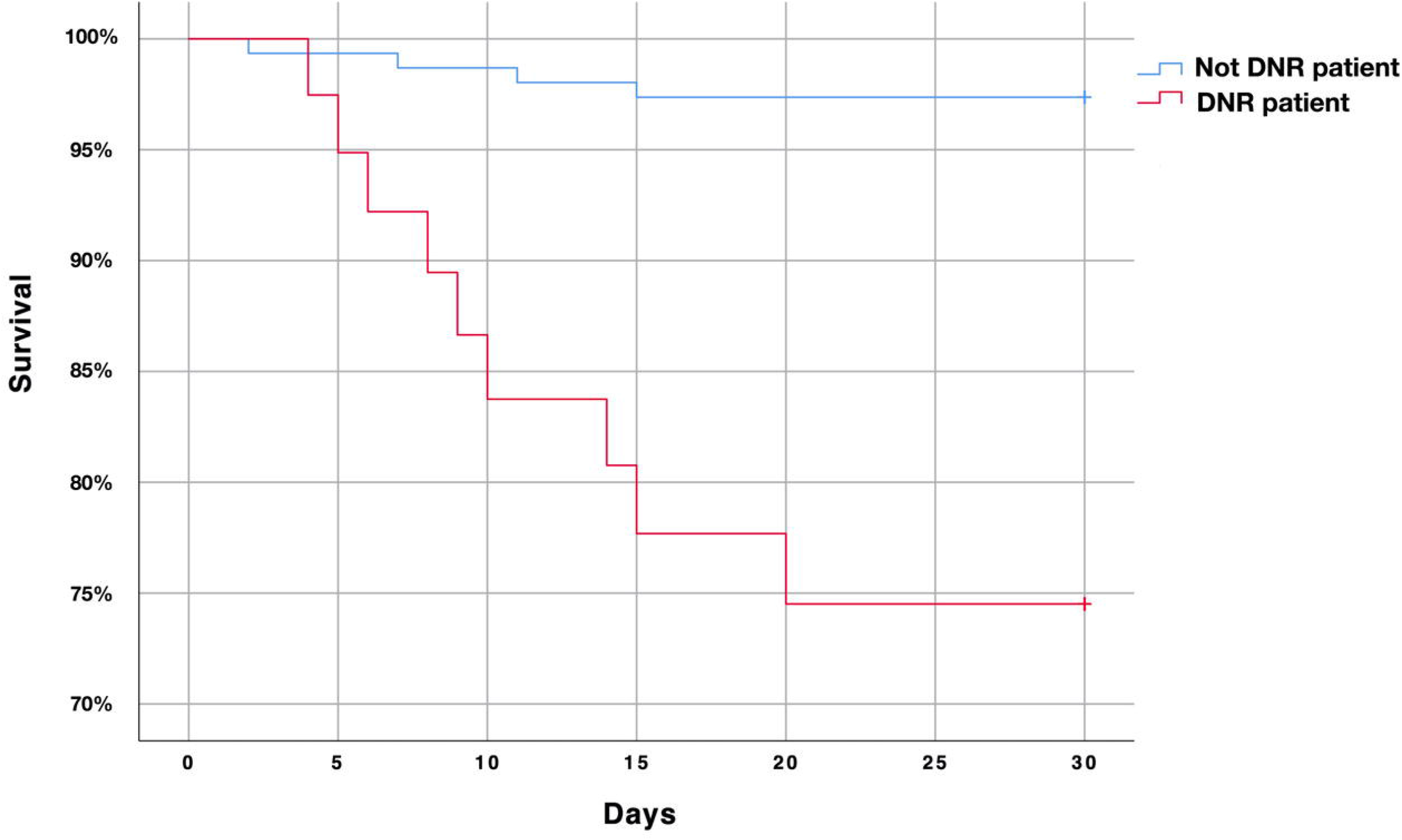
Patients’ survival stratified into DNR/control group, according to standardized national guidelines. As retrospective study, censoring was performed at April 30^th^.

### Secondary Outcomes

Regarding the deceased patients in the *DNR group*, average hospital length-of-stay (LOS) was 6.2 days (3 – 11, SD 2.4); for 8 (89%) of them it was possible to meet a family member before the end of life. All 9 patients (100%) received oxygen therapy until death, 4 (44%) of them had a continuous intravenous fluids infusion and 8 (89%) received at least one antibiotics or anti-retroviral therapy during the entire hospitalization (Table 3); all of them (100%) received opioids (89% morphine, 11% fentanyl), 8 (89%) received sedatives as needed, especially midazolam, while no neuroleptics were administered. Treatment implementation resulted in a significant improvement of RASS-PAL, from a median of 2.2 (SD 1.2) to -1.8 (SD 2), p < 0.0001 (Figure 3), and of the modified Borg scale, from a median of 5.7 (SD 3.3) to 4.7 (SD 3.7), p = 0.581 (Figure 1SM).

**Figure 3.**
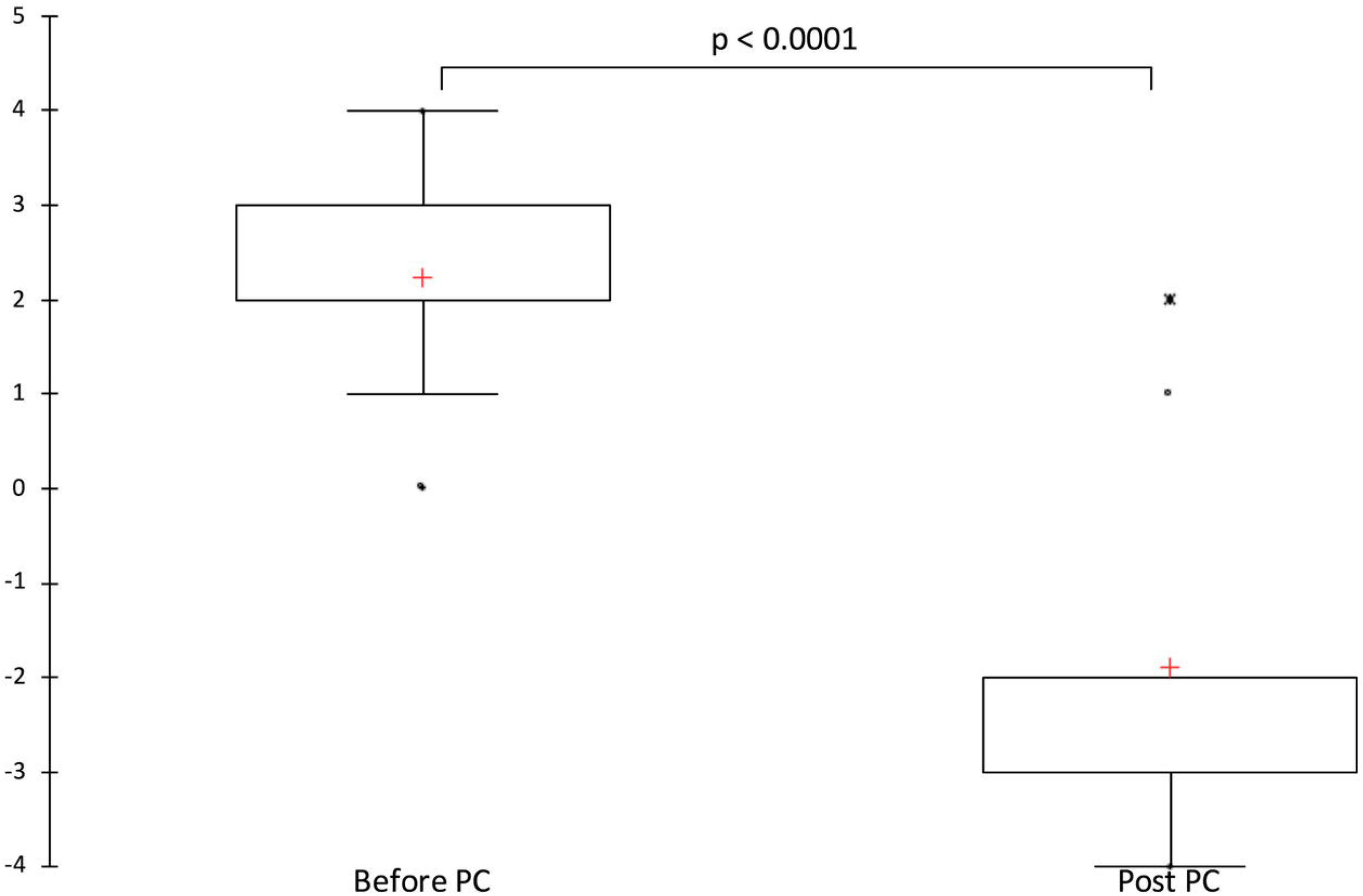
RASS-PAL scale has been used in COVID-19 deceased patients’ agitation assessment. An assessment before and after treatment was performed. After specialistic treatment, RASS-PAL significantly improved from 2.2 to -1.8 (p < 0.0001). Crosses represent the average of the sample.

## DISCUSSION

COVID-19 is a disease characterized by important clinical and social impacts, with a mortality rate that is highest in patients over 60 years ^12^. In this pandemic setting characterized by limited resources, accurately identifying patients at high risk of short-term death (30 days) represents a key point in both patients’ triage and in-hospital management. Application of the Swiss national standardized DNR protocol allowed the identification of COVID-19 patients with a significantly increased short-term mortality rate in comparison to non-DNR COVID-19 patients ^20,21^. This evidence suggests that these guidelines, based on expert opinion and not previously validated, pragmatically identify patients at higher risk of short-term death, thus selecting a population requiring a different specialist approach, rather than intensive care. Unfortunately, DNR definitions change radically between countries, making it difficult to compare these groups of patients. Nevertheless, a national use of shared guidelines made it possible to identify a proportion of patients at high risk of short-term clinical deterioration.

Even though the lack of ICU admission can be considered a bias for the increased mortality in the *DNR group*, these guidelines are able to identify those patients presenting a more rapid deterioration and requiring a more intense clinical support in the short term. A study assessing the *DNR group* patients outcomes in the context of ICU admission could not be performed for obvious ethical and health-costs related reasons, but it can be deduced that the higher rate of clinical worsening seen in DNR patients might correlate with a lack of benefit from ICU admission.

The social impact of COVID-19 led the Public Authority to limit family members’ access to hospitalized patients, in order to preserve Public Health ^22^. In this context, HCP adequate identification of patients at high risk of death allows to predict, and thus to properly organize, a visit permit for patients at the end-of-life. This aspect is essential for Public Health, but also for the long-term social benefits for patients’ relatives, decreasing the risks of post-traumatic-stress-disorder, of unprocessed mourning and of additional psychological diseases^23^. Moreover, it may prevent future increases in health costs, due to the better management of these important psychological and ethical aspects.

Palliative treatment played an important role in taking care of COVID-19 DNR patients, both in terms of psychological and physical symptomatic relief ^22,24,25^. The effectiveness shown in alleviating dyspnea and agitation, even in this relatively small group of DNR patients, underlines the importance of PC in this scenario. The results we obtained with sedation, besides being significant from a statistical point of view, showed us how the clinical handling of these patients has been satisfactory in guarantying them adequate comfort, even in their last moments of life. In order to achieve this important result, and to adequately adapt therapy and management in relation to the clinical status, we realized that it was mandatory to evaluate patient’s agitation/sedation level through the use of appropriate international scales (RASS-PAL in our case) ^18,26^. The assessments carried out so far, in accordance with evidences from other groups ^24^, highlighted the importance of the PC specialist’s role in the better management of symptoms such as agitation, dyspnea, thirst, fear and solitude, which are unfortunately very frequent in the dying population ^25^.

Our study is burdened by some limitations. Firstly, it was a retrospective collection of a cohort of patients, with a small sample of DNR patients; however, the significance level of the statistical tests remains high and should induce other groups to carry out future prospective validations in this regard. The group of DNR patients presented a median age greater than the control group; this may reflect the presence of co-morbidities, a key-elements necessary to identify DNR patients according to national guidelines; however, the increased mortality rate in DNR group was at 30 days, therefore the presence of a median age of 80 years alone was not sufficient to explain this increase in short-term mortality. In the analysis of palliative care treatment we decided to evaluate only the administration of opiates and sedatives and their effects, as a more specific information in this context resulted incomplete due to the retrospective design of the study. Finally, we did not investigate the follow-up in terms of hospital discharge, transfer to other facilities or survival after 30 days, mainly because the primary outcome of our study was to investigate the 30-day overall survival in COVID-19 patients, comparing DNR to not DNR groups.

## CONCLUSIONS

The high mortality rate in COVID-19 patients enhances how it is relevant to identify the subpopulation at a higher risk of short-term death in a pandemic situation characterized by limited resources; in this scenario, the validation analysis we performed supported the role of the Swiss national standardized protocol in DNR patients’ identification. An early access to specialistic treatment should be mandatory, especially for this subgroup of patients, with the aim to improve the end-of-care management and at the same time to reduce the social impact of this pandemics.

## Data Availability

All data are available under adequate written request

