## Supplementary material for "DNR orders in SARS-CoV-2 patients: a retrospective validation study in a Swiss COVID-19 Center": SM

**Title: DNR order in SARS-CoV-2 patients: preliminary guidelines validation and quality of palliative care in a Swiss COVID-19 Center**

**Authors:** Giorgia Lo Presti, MD, Maira Biggiogero, PhD, Andrea Glotta, RN, Carola Biondi, MD, Zsofia Horvath, MD, Rosambra Leo, MD, Alessandra Franzetti-Pellanda, MD, Xavier Capdevila, Prof, Andrea Saporito, PD, Samuele Ceruti, MD

**Address for correspondence:**

Samuele Ceruti, MD, Clinica Luganese Moncucco, Via Moncucco 10, 6900 Lugano

Tel: +41 (0) 91/960.81.08,

**Table 1SM:** Richmond Agitation-Sedation Scale - Palliative version (RASS-PAL)

Physical Stimulation

Verbal Stimulation

| Score | Term | Description |
| --- | --- | --- |
| +4 | **Combative** | Overtly combative, violent, immediate danger to staff (e.g. throwing items); there is a possibility attempting to get out of bed or chair |
| +3 | **Very agitated** | Pulls or removes lines (e.g. IV/SQ/Oxygen tubing) or catheter(s); aggressive, there is a possibility attempting to get out of bed or chair |
| +2 | **Agitated** | Frequent non-purposeful movement, there is a possibility attempting to get out of bed or chair |
| +1 | **Restless** | Occasional non-purposeful movement, but movements not aggressive or vigorous |
| 0 | **Alert and calm** |  |
| -1 | **Drowsy** | Not fully alert, but has sustained awakening (eye-opening/eye contact) to voice (10 seconds or longer) |
| -2 | **Light sedation** | Briefly awakens with eye contact to voice (less than 10 seconds) |
| -3 | **Moderate sedation** | Any movement (eye or body) or eye opening to voice (but no eye contact) |
| -4 | **Deep sedation** | No response to voice, but any movement (eye or body) or eye opening to stimulation by light touch |
| -5 | Not rousable | No response to voice or stimulation by light touch |

**Table 2SM:** Borg modified scale

| Score | Explanation |
| --- | --- |
| 0 | No exertion/breathlessness |
| 0.5 | Very, very light |
| 1 | Very light |
| 2 | Light |
| 3 | Moderate |
| 4 | Somewhat hard |
| 5 | Hard |
| 6 |  |
| 7 | Very hard |
| 8 |  |
| 9 | Very, very hard |
| 10 | Maximal |

**Table 3SM:** Frailty scale

| Frailty Score | Explanation |
| --- | --- |
| 1 – Very Fit | People who are robust, active, energetic and motivated.These people commonly exercise regularly. They are among the fittest for their age |
| 2 – Well | People who have no active disease symptoms but are less fit than category 1. Often, they exercise or are very active occasionally, e.g. seasonally. |
| 3 – Managing Well | People whose medical problems are well controlled, but are not regularly active beyond routine walking. |
| 4 – Vulnerable | While not dependent on others for daily help, often symptoms limit activities. A common complaint is being “slowed up”, and/or being tired during the day. |
| 5 – Mildly Frail | These people often have more evident slowing, and need help in high order IADLs (finances, transportation, heavy housework, medica- tions). Typically, mild frailty progressively impairs shopping and walking outside alone, meal preparation and housework. |
| 6 – Moderately Frail | People need help with all outside activities and with keeping house. Inside, they often have problems with stairs and need help with bathing and might need minimal assistance (cuing, standby) with dressing. |
| 7 – Severely Frail | Completely dependent for personal care, from whatever cause (physical or cognitive). Even so, they seem stable and not at high risk of dying (within ~ 6 months). |
| 8 – Very Severely Frail | Completely dependent, approaching the end of life.Typically, they could not recover even from a minor illness. |
| 9 – Terminally Ill | Approaching the end of life. This category applies to people with a life expectancy <6 months, who are not other ise evidently frail. |

**TABLE 4SM Chi-square analysis**

|  | No | Yes |  |
| --- | --- | --- | --- |
| Alive | 169 | 31 | 200 |
| Dead | 4 | 9 | 13 |
|  | 173 | 40 | 213 |

|  | Value | P value (2-sided) |
| --- | --- | --- |
| Pearson Chi-Square | 23.104 | < 0.0001 |
| Continuity Correction | 19.716 | < 0.0001 |
| N of Valid cases | 213 |  |

*Chi-square analysis for patients on PC versus not on PC patients, compared to 30-day survival. The test rejects the hypothesis that the distribution of patients with respect to belonging to the PC group is random, identifying instead a clear existing correlation.*

**Figure 1 SM**

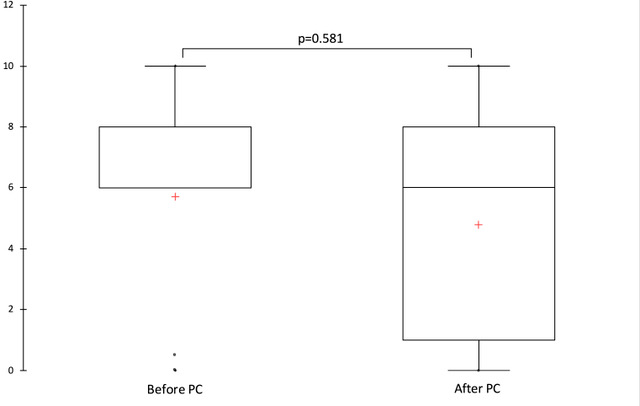

*Modified Borg scale has been used in COVID-19 deceased patients’ dyspnea assessment. An assessment before and after PC treatment was performed. After PC treatment, dyspnea improved from 5.7 to 4.7 (p = 0.29). Crosses represent the average of the sample.*
